# Quality of Labour and Delivery Care Process and Associated Factors in Government Hospitals of Ethiopia: a multilevel analysis

**DOI:** 10.1101/2023.04.14.23288601

**Authors:** Negalign B Bayou, Liz Grant, Simon C Riley, Elizabeth H Bradley

**Author notes:** These authors contributed equally to this work. Corresponding author (NBB).

## Abstract

**Background:** Ethiopia has one of the highest maternal mortality ratios in Africa. Few have examined the quality of labour and delivery (L&D) care in the country. This study evaluated the quality of routine L&D care and identified patient-and hospital-level factors associated with the quality of care in a subset of government hospitals.

**Materials and methods:** This was a facility-based, cross-sectional study using direct non-participant observation carried out in 2016. All mothers who received routine L&D care services at government hospitals (n=20) in one of the populous regions of Ethiopia, Southern Nations Nationalities and People’s Region (SNNPR), were included. Mixed effects multilevel linear regression modeling was employed in two stages using hospital as a random effect, with quality of L&D care as the outcome and selected patient and hospital characteristics as independent variables. Patient characteristics included woman’s age, number of previous births, number of skilled attendants involved in care process, and presence of any danger sign in current pregnancy.

Hospital characteristics included teaching hospital status, mean number of attended births in the previous year, number of fulltime skilled attendants in the L&D ward, whether the hospital had offered refresher training on L&D care in the previous 12 months, and the extent of resources available (measured on a 0-100% scale) to provide quality L&D care as defined by the Ethiopian Ministry of Health in 2014. The outcome was measured with a quality of L&D care score (scale 0 to 100) based on adherence to L&D care standards, which had been introduced by the Ethiopian Ministry of Health in 2014.

**Results:** On average, the hospitals met two-thirds of the standards for L&D care quality, with substantial variation between hospitals (standard deviation 10.9 percentage points). While the highest performing hospital met 91.3% of standards, the lowest performing hospital met only 35.8% of the standards. Hospitals had the highest adherence to standards in the domain of immediate and essential newborn care practices (86.8%), followed by the domain of care during the second and third stages of labour (77.9%). Hospitals scored substantially lower in the domains of active management of third stage of labour (AMTSL) (42.2%), interpersonal communication (47.2%), and initial assessment of the woman in labour (59.6%). We found the quality of L&D care score was significantly higher for women who had a history of any danger sign (β = 5.66; p-value = 0.001) and for women who were cared for at a teaching hospital (β = 12.10; p-value = 0.005). Additionally, hospitals with lower volume and more resources available for L&D care (P-values < 0.01) had higher L&D quality scores.

**Conclusions:** Overall, the quality of L&D care provided to labouring mothers at government hospitals in SNNPR was limited. Lack of adherence to standards in the areas of the critical tasks of initial assessment, AMTSL, interpersonal communication during L&D, and respect for women’s preferences are especially concerning. Without greater attention to the quality of L&D care, regardless of how accessible hospital L&D care becomes, maternal and neonatal mortality rates are unlikely to decrease substantially.

## Background

Globally, more than 300,000 mothers die annually due to preventable pregnancy and childbirth related problems (1). Almost all (99%) of these deaths occur in low-and middle-income countries (LMICs) (2), of which more than two thirds were in sub-Saharan Africa (SSA). Ethiopia has one of the highest maternal mortality ratios (MMR) in SSA, 412 per 100,000 live births (3). While countries in SSA have experienced substantial reductions in MMR largely due to increased access to skilled birth attendance (4) in the last thirty year, MMR in SSA continues to lag far behind. Ethiopia is not on track to meet Sustainable Development Goals of reducing MMR to less than 70 per 100,000 live births and ending preventable deaths of newborns to at least as low as 12 per 1,000 live births by 2030 (5, 6).

While access to skilled birth attendance is essential, newer evidence is emerging on the importance of enhancing the quality of labour and delivery (L&D) care in order to reduce MMR. Half of all maternal deaths and one million newborn deaths can be prevented annually by providing high-quality care before, during, and after childbirth (7–11). Furthermore, qualitative data from Ethiopia (12) have suggested that perceptions of poor quality of hospital-based L&D care can be a major barrier in scaling up the use of available skilled birth attendants, limiting their impact on MMR.

Although researchers and policymakers have underscored the importance of quality of L&D care (7), few empirical studies in SSA have examined the quality of L&D services, and those that have (13–15) generally described overall quality without examining the patient- and hospital-level factors that were associated with poor quality of L&D services. The one study we found that used a substantial sample of hospitals to examine factors associated with L&D quality was from Rwanda (12) and is also more than a decade old; this study (12) found no significant patterns in quality of L&D care by hospital-level factors. Thus, the contemporary level of quality of hospital-based L&D care as well as the patient- and hospital-level factors associated with higher L&D care quality in SSA in general and in Ethiopia, specifically, remain largely unknown.

Accordingly, we sought to describe the quality of hospital-based L&D care as measured by adherence to L&D quality of care standards set forth by the Ethiopian Ministry of Health (16) and to identify patient- and hospital-factors associated with higher quality L&D hospital care in Ethiopia. We used a non-participant observation study conducted in 20 government hospital in the most populous region of Ethiopia, Southern Nations Nationalities and People’s Region (SNNPR). Findings may be useful for designing and targeting intervention to improve quality of care and hence reduce MMR.

## Materials and methods

### Study design

We undertook a non-participant observation study conducted with all normal vaginal deliveries of women 18 years and older occurring in government hospitals (n=20) in the Ethiopian region of SNNPR from November 11 to December 10, 2016. Births were excluded (n=16 births) if the birthing woman had any high-risk factors (e.g., pre-eclampsia or previous scar), if the neonate was not alive at birth, or if the birthing woman developed complications during the during the first hour following the birth.

### Data collection

In each hospital, non-participant observation was undertaken by two experienced, professional midwives identified as outside assessors and previously not known to the hospital staff. The birthing process for women who were attended between 8:30am and 12:30pm and between 1:30pm and 5:30pm and between 6:30pm and 9:30pm were observed until one hour postpartum. In high volume hospitals, data collectors observed the maximum number of births possible without compromising data quality. If only one woman was in active labour, both data collectors observed. If two women who were in active labour were being attended simultaneously, the data collectors observed one each, and if more than two were occurring simultaneously, the data collectors selected two women using a lottery method and observed one each.

Using an observation checklist, data collectors assigned score of ‘1’ if the activity were observed and performed according to the recommended standard of care and ‘0’ otherwise. At the end of a completed observation and before starting a new one, data collectors reviewed the clinical record of the woman to gather data about her initial assessments based on the history and physical examination completed before the admission observation began.

### Measures

#### Dependent variable

The outcome was calculated for each birth as the percentage of all relevant standards fulfilled, with possible values from 0%-100%. The percentage fulfilled was reported for the relevant standards overall as well as within 7 domains. For each hospital, the quality of L&D care score was determined as the mean percentage across all observed births in that hospital, reported for the standards overall and within each of 7 domains.

#### Independent variables

Patient-level independent variables included the woman’s age, number of previous births, number of skilled attendants involved in care process, presence of any danger sign in current pregnancy. Hospital-level independent variables included teaching hospital status, mean number of births in the hospital in the previous year, number of fulltime skilled attendants in the L&D ward on the assessment day, whether the hospital had offered refresher training on L&D care in the previous 12 months, and the extent of resources available (measured on a 0-100% scale) to provide quality L&D care as defined by the Ethiopian Ministry of Health (16). Details on the measure of resources available have been previously described (17).

### Data analysis

We used descriptive statistics to summarize both the outcome (overall and for 7 domains) and the patient- and hospitals-level independent variables. We examined the unadjusted associations between the outcome and each independent variable using t-tests for binary and categorical independent variables and Pearson correlation coefficients for continuous independent variables. Results available from first author.

Multivariable regression was employed to estimate adjusted associations between hospital quality of L&D care scores and the independent variables. Mixed effects multilevel linear regression was fitted in two stages to achieve a parsimonious predictive model. The first stage, or null model, was fitted with no explanatory variables using hospital as a random effect. This model was used to determine how being attended in a particular hospital may explain differences in the outcome. The second stage, or the full model, included the patient-and hospital-level independent variables.

Intra-class correlation coefficients (ICCs) were also calculated for both the null and full models to explore the amount of variance in the outcome attributable to differences within versus between hospitals. Because of the centering of all continuous independent variables on the grand means, the random term of the intercept at hospital level adjusted for the variance in outcome attributable to the hospital, and appropriately controlled for the clustering of observations by hospital. Interaction terms were created, and their effects tested. Cases with missing data for key variables were excluded from further analysis. P-value <0.05 was used to determine statistical significance of the associations. Data analysis was accomplished using the Advanced Statistics Module of SPSS IBM Statistics.

Ethics approval was obtained from the Institutional Review Boards (IRBs) of Jimma University (Ethiopia) and The University of Edinburgh (UK). All participants provided informed consent following the procedure approved by the IRBs before being observed. The authors did not have access to information that could identify individual participants during or after data collection.

## Results

### Characteristics of the study participants and hospitals

A total of 1,351 labouring mothers were approached and gave consent to participate in the study; 16 were later excluded due to non-normal progression of labour resulting in an analytic sample of 1,335 women (98.8% of those approached for the study).

The mean age of women in the study was 25.7 years (standard deviation (SD) = 4.8 years), and about two thirds were aged 21-30 years **(Table 1)**. Nearly three quarters (74.2%) had previously given birth to one or two children. More than three quarters (77.1%) of the women had had 3 or 4 antenatal care (ANC) visits for the present pregnancy. Of the 1,335 women in the analytic sample, 38 women (2.8%) encountered one or more danger signs in the present pregnancy. Just more than half (57.8%) of the women were attended at a non-teaching type of hospital. The baby was alive and in good condition (with no complication) until the end of observation in 95.6% (n = 1,275) of the cases, while the rest (n = 58) were alive with complication **(Table 1)**.

**Table 1:** Characteristics of women observed (N=1,335) for quality of L&D care at public hospitals in SNNPR, Ethiopia, 2016

| Characteristics | n | % |
| --- | --- | --- |
| Age of women |  |  |
| 20 and below | 244 | 18.4 |
| 21-25 | 487 | 36.7 |
| 26-30 | 441 | 33.3 |
| 31-35 | 119 | 9.0 |
| 36-40 | 40 | 3.0 |
| 41 and above | 4 | 0.3 |
| Missing | 9 | 0.7 |
| Number of previous pregnancies |  |  |
| 1-2 | 754 | 66.8 |
| 3-4 | 269 | 23.8 |
| 5 and above | 106 | 9.4 |
| Missing | 206 | 15.4 |
| Number of previous births |  |  |
| 0 | 87 | 6.5 |
| 1-2 | 808 | 74.2 |
| 3-4 | 201 | 18.5 |
| 5 and above | 80 | 7.3 |
| Missing | 159 | 11.9 |
| Number of ANC visits in current pregnancy |  |  |
| 1-2 | 246 | 20.4 |
| 3-4 | 929 | 77.1 |
| 5 and above | 30 | 2.5 |
| Missing | 130 | 9.7 |
| Encountered any danger sign in current pregnancy |  |  |
| Yes | 38 | 2.8 |
| No | 1297 | 97.8 |
| Position of the woman during childbirth |  |  |
| Supine (lithotomy) | 1319 | 98.8 |
| Lateral tilt | 16 | 1.2 |
| Foetal outcome at the end of observation |  |  |
| Alive with no complication | 1275 | 95.6 |
| Alive with complication | 58 | 4.4 |
| Missing | 2 | 0.5 |

A total of 35% of the hospitals were teaching hospitals **(Table 1A)**. On average, hospitals had 49,077 births in the previous year, with a mean service volume of 2454 deliveries (SD 1293.3) in a hospital. Hospitals had on average 440 fulltime skilled attendants in the L&D ward, with an average of 22 (SD 9.7) per hospital. A total of 45% of the hospitals had offered refresher training on management of L&D in the past 12 months. The average score (17) assessing the extent of resources available to provide quality of L&D care was 68.2%, with a SD of 15.9 percentage points.

**Table 1A:** Characteristics of hospitals (N=20)

| Characteristics | N (or mean percentage score) | % (or SD) |
| --- | --- | --- |
| Teaching status |  |  |
| Teaching | 7 | 35 |
| Non-teaching | 13 | 65 |
| Number of fulltime skilled attendants in labour ward | 22.0 | 9.7 |
| Refresher training on L&D care given in previous 12 months |  |  |
| Yes | 9 | 45 |
| No | 11 | 55 |
| Number of births attended in last year | 2454.0 | 1293.3 |
| Measure of resources available (0-100% scale) | 68.2 | 15.9 |

### Outcome

#### Summary score for quality of L&D care

The mean percentage of standards fulfilled across all hospitals was 66.6% (standard deviation [SD] 10.9 percentage points) **(Fig 1)**. The highest performing hospitals met 91.3% of standards; the lowest performing hospital met 35.8% of the standards. Hospitals achieved the highest mean percentage score in the domain of immediate and essential newborn care practices (86.8%, SD 11.8 percentage points), followed by the domain of second and third stages of labour 77.9% (SD 20.6 percentage points). The domain with the lowest mean percentage score was in complying with AMTSL standards (42.2%, SD 12.2 percentage points), followed by the domains of interpersonal communication (47.2%, SD 38.4 percentage points) and initial assessment of the woman in labour (59.6%, SD 15.6 percentage points).

**Fig 1:**
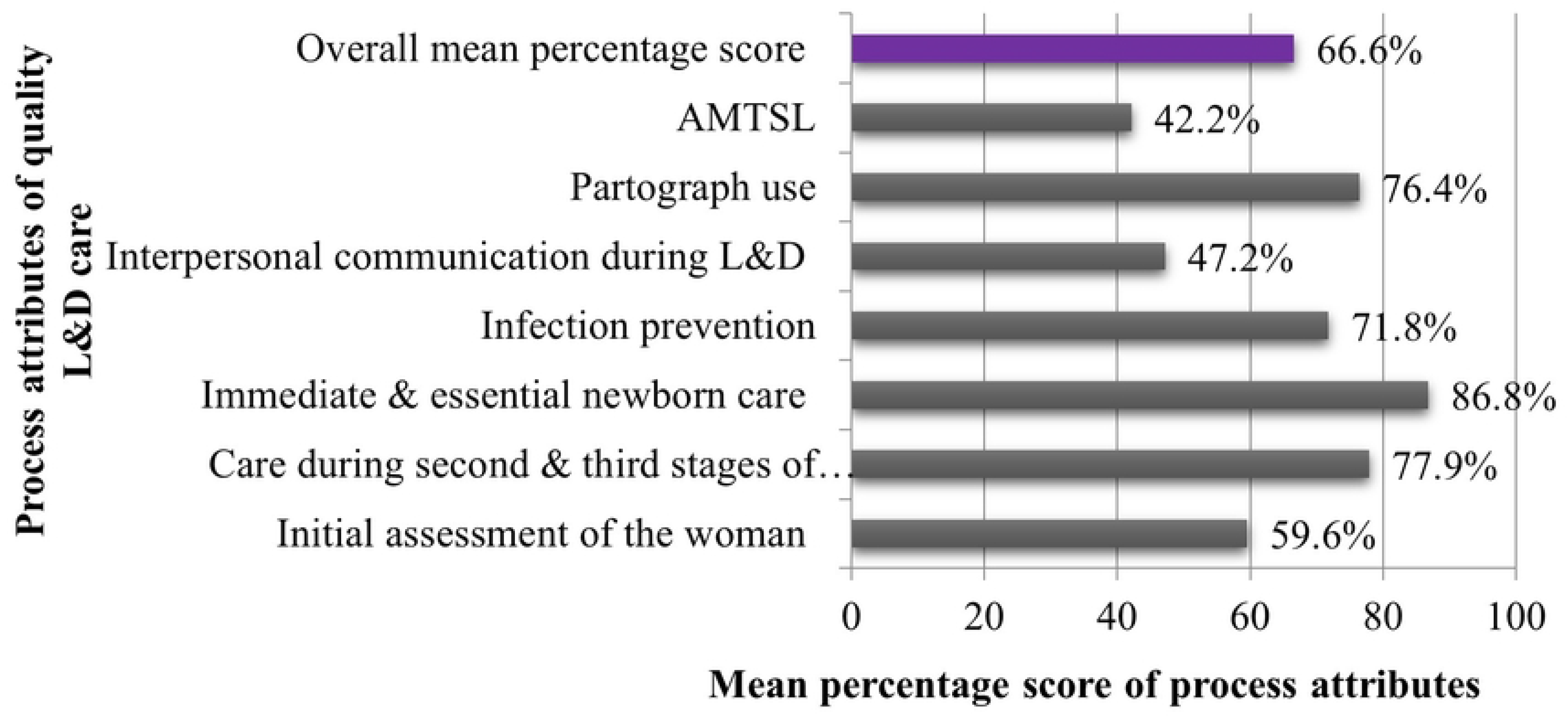
Summary of quality of L&D care, overall and by domain, at public hospitals in SNNPR, Ethiopia, 2016

#### Domain 1: Initial assessment

Overall, providers adhered to an average of 59.6% of the required initial assessment standards, SD 15.6 percentage points. The mean percentage scores varied substantially across hospitals with the highest fulfilling 74.8% of the standards (SD 12.7) and the lowest fulfilling 30.6% of the standards (SD 15.1). The most commonly documented aspects of the initial assessment were vaginal examination findings, (95.3%), which included checking and recording cervical dilatation, foetal presentation, and membrane status and pulse rate (93.8%). In contrast, documentation of findings from the general examination was generally less common (58.4%). The woman’s temperature was recorded for about half (52.5%) of the women. The previous obstetric history of complication during previous pregnancies was recorded for 6% of the women. Similarly, providers recorded whether the woman had any danger sign in the present pregnancy in only 2.8% of the cases.

#### Domain 2: Second and third stages of labour

In this domain, providers adhered to an average 77.9% of the recommended standards, SD 20.6 percentage points. The mean percentage scores were as high as 99.1% (SD 4.1 percentage points) in one hospital and as low as percentage points54.0% (SD 21.2 percentage points) for another hospital. Scores were highest (92.1%–95.1%) for several tasks including examination of the placenta and membranes for completeness, palpation of the uterus every 15 minutes after the placenta is delivered to confirm uterine contraction, and examination of the vulval-perineal region for possible lacerations. In contrast, scores were far lower for other tasks. For example, checking for vaginal bleeding and taking blood pressure and pulse rate every 15 minutes after birth were performed in just about half (52.8%) and less than one third (30.5%) of the cases, respectively (for more information on this domain, see **Table 2)**.

**Table 2:** Standards fulfilled in the second and third stages of labour, and immediate and essential newborn care in the first hour postpartum at public hospitals in SNNPR, Ethiopia, 2016

| Domain of care process | Observed deliveries (N=1335) |  |
| --- | --- | --- |
|  | n | % |
| **Care during the second and third stages of labour (mean percentage score = 72.9% [SD 20.6])** |  |  |
| Examined completeness of placenta and membranes | 1221 | 92.1 |
| Palpated uterus after placenta is delivered to confirm uterine contraction | 1063 | 79.6 |
| Examined vulval-perineal region for possible lacerations | 1229 | 93.0 |
| Checked vaginal bleeding every 15 minutes after placenta is delivered | 686 | 52.8 |
| Checked mother's vital sign (pulse rate & BP) every 15 minutes | 406 | 30.5 |
| **Immediate and essential newborn care practice (mean percentage score = 86.8% [SD 11.8])** |  |  |
| Delivered to mother's abdomen | 1170 | 87.6 |
| Immediately dried with warm towel | 1289 | 96.6 |
| Wiped eyes as s/he stimulated breathing | 1255 | 94.0 |
| Cut cord, tied 2 fingers from abdomen & another 2 fingers from the first | 1195 | 89.5 |
| Placed skin to skin on mother's chest or abdomen for at least 30 minutes to initiate breastfeeding | 498 | 37.6 |
| Applied tetracycline eye ointment to both eyes | 1163 | 87.6 |
| Gave vitamin K 1mg IM | 1214 | 91.3 |
| Weighed and classified | 1312 | 98.7 |

#### Domain 3: Immediate and essential newborn care

Overall, provides complied to an average 86.8% of the standards for immediate and essential newborn care practices, SD 11.8 percentage points **(Table 2)**. Providers performed the recommended tasks in 87.6% to 98.7% of the deliveries, and the performance was less variable across hospitals relative to other domains with most hospitals having a mean percentage score above 80%. The vast majority (98.7%) of newborns were weighed and classified and 96.6% were dried with a towel immediately after birth; however, immediate post-partum care was often lacking. For instance, providers helped the mother initiate breastfeeding within the first hour post-partum in 37.6% of the cases, checked the newborn’s temperature every 15 minutes in only 21.9% of the cases, and explained the importance of delayed bathing for the first 24 hours after birth to just more than half (52.3%) of the women.

#### Domain 4: Infection prevention (IP) practice

Providers adhered to an average 71.8% of the recommended IP standards, SD 16.6 percentage points. The scores varied across hospitals, ranging between 90.5% (SD 7.9 percentage points) to 41.3% (SD 11.8 percentage points). Providers correctly wore gloves, disposed sharps and decontaminated instruments and surfaces after procedure most of the time (96.7%, 91.8% and 96.5%, respectively); however, providers rarely wore other protective clothing other than gloves (14.4%), and hand hygiene before examination during labour occurred in only 38.8% of the cases (for more information from this domain, see **Table 3)**.

**Table 3:** IP practices during routine L&D care and partograph use at public hospitals in SNNPR, Ethiopia, 2016

| Domain of care process | Observed deliveries (N=1335) |  |
| --- | --- | --- |
|  | n | % |
| **IP practices (mean percentage score = 71.8% [SD 16.6])** |  |  |
| Washed hands before examination during labour | 503 | 38.8 |
| Put on two pairs of sterile gloves for vaginal examination | 1291 | 96.7 |
| Put on clothing to protect face and other parts of the body | 192 | 14.4 |
| Decontaminated all reusable instruments in 0.5% chlorine solution | 1226 | 91.8 |
| Disposed all sharps in a yellow-colored puncture proof container immediately after use | 1284 | 96.5 |
| Disposed all contaminated wastes in a red-colored leak proof container<sup>1</sup> | 1038 | 77.9 |
| Disposed non contaminated materials in a container designated for general waste | 1063 | 79.7 |
| Washed hands thoroughly with soap and water, and dried them | 1015 | 76.4 |
| **Partograph use (mean percentage score = 76.3% [SD 23.9])** |  |  |
| Progress of the labour monitored using partograph | 1073 | 80.4 |
| While the woman was in active labor, plotted the following physical exam measurements on partograph (n=1073): |  |  |
| FHB at least every half hour | 918 | 85.6 |
| Cervical dilatation at least every 4 hours | 975 | 91.2 |
| Descent of head at least every 4 hours | 841 | 78.7 |
| Frequency & duration of contractions at least every half hour | 902 | 84.4 |
| Blood pressure at least every 4 hours | 866 | 81.1 |
| Temperature at least every 2 hours | 390 | 36.6 |

<sup>1</sup>Contaminated wastes included for example gloves and swabs.

#### Domain 5: Use of partograph to monitor labour

The overall mean percentage score for correct use of partograph was 76.4%, SD 23.9 percentage points. Hospitals varied substantially in this area of practice with providers at one hospital correctly used partograph all the time (100%), while those at another doing so in about half (52.1%, SD 19.4 percentage points) of the cases. In cases for which providers used a partograph to monitor the progress of labour (n=1073), they plotted cervical dilatation on the partograph at least every 4 hours in 91.2% of the cases, but providers recorded temperature at least every 2 hours in only 36.6% of the cases (for more information from this domain, see **Table 3)**.

#### Domain 6: Active management of third stage of labour (AMTSL)

Providers met an average 42.4% of the AMTSL standards, SD 12.2 percentage points. Three hospitals performed the highest in this area, scoring 50.5% in this domain (SD 0 – 0.2 percentage points), while the lowest performing hospital scored 24.5% (SD 10.9 percentage points). Although 97.2% of the women received a uterotonic, i.e., oxytocin or ergometrine, and almost all of them received it within three minutes of delivery, and 90.8% within one minute of delivery, controlled cord traction was performed in only 74.1% of cases, and uterine massage was performed in only 42.4% of the observed deliveries.

#### Domain 7: Interpersonal communication during L&D

Providers adhered to an average 47.2% of the recommended practices of interpersonal communication during L&D, SD 38.4 percentage points. Three hospitals scored above 82% (SD ranging between 10.7 and 32.8 percentage points), while another three scored below 15% (SD 13.2 – 34.2 percentage points). Providers gave explanations of procedures and what would happen during labour in about two thirds (63.1%) of the labours observed, but providers encouraged only about a third (31.4%) of the women to have a support person present throughout L&D.

### Factors associated with quality of L&D care process

In the fully adjusted model with hospital as a random effect (ICC in null model 0.59), both patient and hospital characteristics were significantly associated with quality of L&D care **(Table 4)**. Among the patient characteristics examined, the presence of a danger sign in the current pregnancy was significantly (and positively) associated with quality of L&D care (β = 5.66; P-value = 0.001). None of the other patient characteristics examined—age of the woman, number of previous births, or the number of skilled attendants involved in care processes—was significantly associated with quality of L&D care. Among hospital characteristics examined, type of hospital (teaching versus non-teaching), lower hospital volume of births in the previous year, and having more resources available for L&D care were each associated with higher quality of L&D care (P-values < 0.01). Neither the number of fulltime skilled health professionals working in the maternity ward nor provision of refresher training on management of L&D in the past 12 months were significantly associated with quality of L&D care (P-values > 0.05).

**Table 4:**
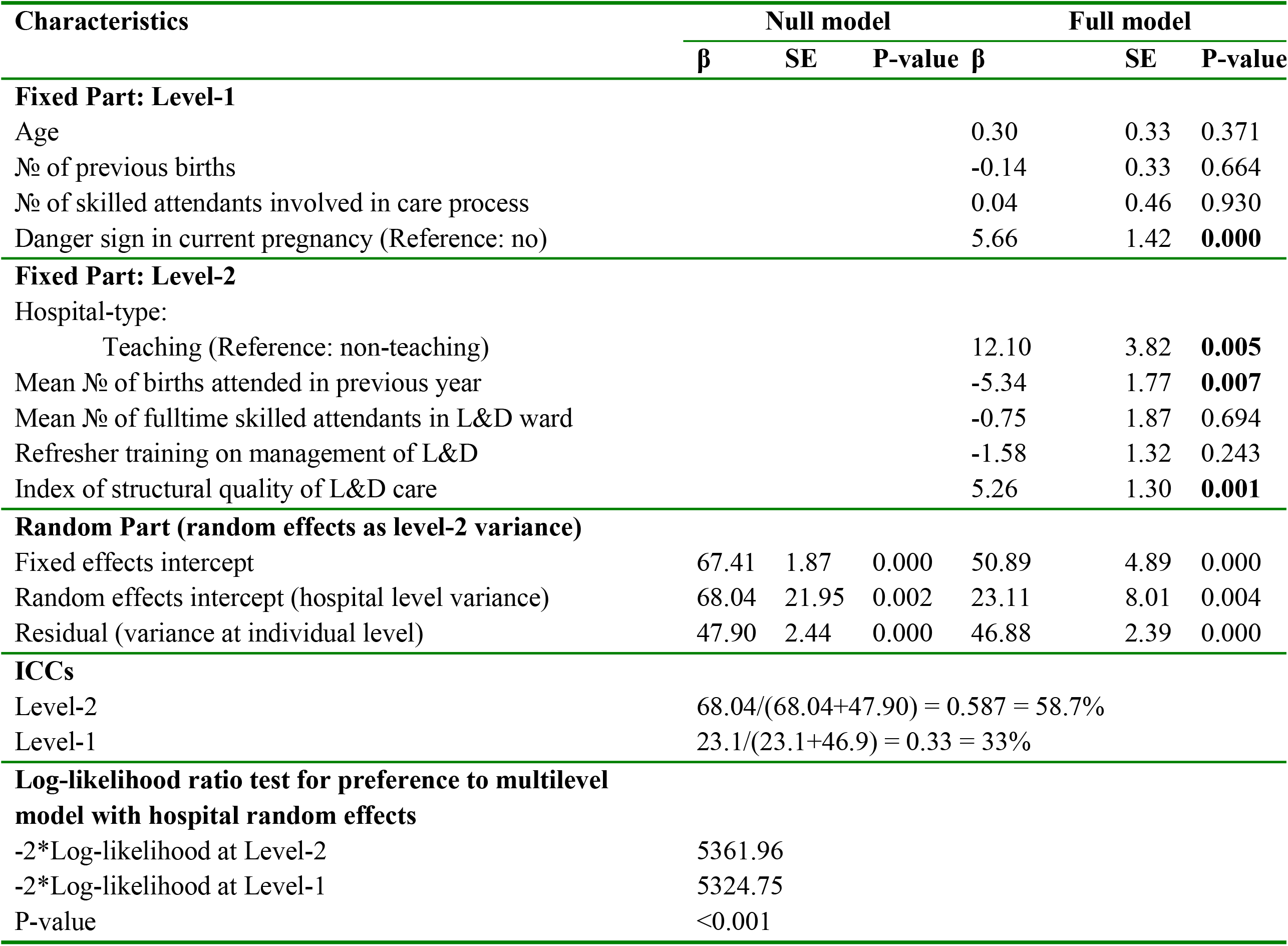
Predictors of the index of quality of L&D care provision at public hospitals in SNNPR, Ethiopia, 2016

| Characteristics | Null model |  |  | Full model |  |  |
| --- | --- | --- | --- | --- | --- | --- |
|  | β | SE | P-value | β | SE | P-value |
| **Fixed Part: Level-1** |  |  |  |  |  |  |
| Age |  |  |  | 0.30 | 0.33 | 0.371 |
| No of previous births |  |  |  | -0.14 | 0.33 | 0.664 |
| No of skilled attendants involved in care process |  |  |  | 0.04 | 0.46 | 0.930 |
| Danger sign in current pregnancy (Reference: no) |  |  |  | 5.66 | 1.42 | **0.000** |
| **Fixed Part: Level-2** |  |  |  |  |  |  |
| Hospital-type: |  |  |  |  |  |  |
| Teaching (Reference: non-teaching) |  |  |  | 12.10 | 3.82 | **0.005** |
| Mean No of births attended in previous year |  |  |  | -5.34 | 1.77 | **0.007** |
| Mean No of fulltime skilled attendants in L&D ward |  |  |  | -0.75 | 1.87 | 0.694 |
| Refresher training on management of L&D |  |  |  | -1.58 | 1.32 | 0.243 |
| Index of structural quality of L&D care |  |  |  | 5.26 | 1.30 | **0.001** |
| **Random Part (random effects as level-2 variance)** |  |  |  |  |  |  |
| Fixed effects intercept | 67.41 | 1.87 | 0.000 | 50.89 | 4.89 | 0.000 |
| Random effects intercept (hospital level variance) | 68.04 | 21.95 | 0.002 | 23.11 | 8.01 | 0.004 |
| Residual (variance at individual level) | 47.90 | 2.44 | 0.000 | 46.88 | 2.39 | 0.000 |
| **ICCs** |  |  |  |  |  |  |
| Level-2 | 68.04/(68.04+47.90) = 0.587 = 58.7% |  |  |  |  |  |
| Level-1 | 23.1/(23.1+46.9) = 0.33 = 33% |  |  |  |  |  |
| **Log-likelihood ratio test for preference to multilevel model with hospital random effects** |  |  |  |  |  |  |
| -2*Log-likelihood at Level-2 | 5361.96 |  |  |  |  |  |
| -2*Log-likelihood at Level-1 | 5324.75 |  |  |  |  |  |
| P-value | <0.001 |  |  |  |  |  |

## Discussion

We found that on average hospitals met only two-thirds of the standards for quality of L&D care, with particularly poor performance in three domains of care: initial assessment of the woman in labour, AMTSL, and interpersonal communication. Importantly, although quality of care was limited overall, one hospital achieved more than 90% of standards met, providing an example of “positive deviance” (18) that could yield information and inspiration for future quality improvement efforts among hospitals in SNNPR. The study also revealed that both patient and hospital characteristics were associated with the quality of L&D care.

The finding on the initial assessment tasks, completed in about 60% of the cases, was lower than previously reported in studies from other regions of Ethiopia (19, 20). For instance, Getachew and colleagues in a USAID-funded study (19 with far less direct observation of births reported that providers performed these tasks in 80–100% of the cases; and Yigzaw and colleagues in a study of Amhara region hospitals reported that providers performed these tasks in 73.6% of the cases (20). The differences area likely due to differences in methodology and region sampled; regardless, the present study highlights the prevalence of missed opportunities for timely identification of women in need of further care or referral. The high-performance levels on examination of the placenta, membranes and vulval-perineal region, and uterine massage yet low performance on monitoring the condition of a mother with vaginal bleeding are findings that are consistent with other studies conducted in SSA (20). Lack of monitoring of vaginal bleeding signifies the potential risk of developing immediate post-partum complications endangering the mother’s life.

Several studies have similarly reported low performance on putting newborn skin-to-skin with the mother (19, 21, 22). Low score for advising about delaying baby bath is also consistent with a similar study (13). Thus, enhancing compliance to the related standards is imperative. The low hand washing practice observed in this study is higher than a study conducted in Ethiopia six years ago (19) and comparable with another study conducted in Malawi (23); it is also consistent with several studies conducted in low-income settings (19, 24, 25), and appears to be an important gap as it risks maternal sepsis and cross infections with fatal diseases like HIV and hepatitis B (19).

Consistent use of partograph for labour monitoring is much better than for previous studies conducted in different countries including, Uganda (26), and Ethiopia (19), and almost comparable with a study conducted in Kenya (27). When correctly used to monitor progress of labour, a partograph can help improve fetal and newborn survival, and significantly reduces unnecessary interventions (19, 28). It is important to explore the reasons for non-use and incorrect use of this effective intervention. The finding on compliance to all AMTSL tasks is also much higher than the study conducted in Ethiopia which reported 29% (19). This may partly be explained by larger sample size used in the present study as well as the time gap between the studies (2010 versus 2016); the current finding may reflect the result of the Government’s efforts towards safe motherhood such as massive human resource and health facility development strategies.

The inadequate performance on interpersonal communication and respecting women’s preferences is consistent with previous studies (19, 22, 29). Although some have argued (30) that such communication may be unrecognized and thus underestimated by data collectors, our findings nonetheless highlight potential opportunities for improvement as poor interpersonal communication and lack of responsiveness to preferences can limit maternal health services utilization (31).

The negative correlation between service volume and quality of care is consistent with previous studies conducted in low-income settings (22, 32). Low volume hospitals may tend to pay more attention to quality of care than hospitals that are overwhelmed with patient volume. Additionally, observing higher quality of L&D care at teaching versus non-teaching hospitals highlights the important role of senior attending professionals, and is consistent with the premise that high quality providers are critical to produce high quality outcomes (33).

The findings should be interpreted in light of several limitations. First, composite scores reflect averages that can mask meaningful variation in individual item responses (34), although they can be more reliable than individual items (35, 36). We have provided ranges and standard deviations to augment the composite score summary statistics. Summary scores can also facilitate comparison and simplify communication of performance on technical components of care to non-technical audiences (35).

Second, direct observation can influence measures of performance due to the inevitable Hawthorne effect (34, 37); however, we sought to minimize this bias by observing hospital staff for a full month and assured anonymity in all data reports. Third, the sample was restricted to hospitals from SNNPR; results may differ in other geographies. Finally, we were unable to collect information on education and income of the women, factors which may influence quality of L&D care. Future studies are warranted to further explore the role of socioeconomics factors in quality of L&D care received.

## Conclusions

In sum, the study indicates substantial opportunities for improvement in the quality of routine L&D care provided both to the women and their babies at Government hospitals in Ethiopia. In particular, screening and monitoring of maternal and newborn conditions were inadequate. Also, early initiation of breastfeeding by placing skin-to-skin with mother, prevention of hypothermia, and advice on delayed baby bath for the first 24 hours were implemented less often than recommended, and critical tasks including hand washing, use of a partograph, AMTSL, and interpersonal communication during L&D were limited. Quality of L&D care was most concerning in high volume, non-teaching hospitals.

At the same time, our study identified hospitals that demonstrated positive deviance (18) in the sense that they performed at high levels despite the challenges all hospitals faced. For instance, in one hospital, the percentage of standards adhered to exceeded 90%. Future studies may examine such institutions more deeply to understand how to achieve and sustain high quality of L&D care, and more importantly, how to spread such practices to other hospitals in need of improvement.

## Data Availability

All relevant data are within the manuscript.

## List of abbreviations

AMTSL: Active management of third stage of labour
FHB: Fetal heart beat
ICCs: Intra-class Correlation Coefficients
IP: Infection prevention
L&D: Labour and delivery
LMICs: Low-and middle-income countries
MMR: Maternal mortality ratio
SNNPR: Southern Nations Nationalities and People’s Region
SSA: Sub-Saharan Africa

## Acknowledgements

We would like to thank SNNPR Regional Health Bureau and administrators of the hospitals for facilitating the data collection process. We are also grateful to the study participants and field research team for their contribution to collecting the data, and Jimma University for offering office space and facilities to ensure data security.

## References

1. WHO, UNICEF, UNFPA, World Bank Group, United Nations Population Division. Trends in maternal mortality: 1990-2015. Geneva: World Health Organization, 2015.

2. Say, L., et al. Global causes of maternal death: a WHO systematic analysis. The Lancet Global Health, 2014, Vol. 2. pp.e323–e333.

3. CSA Ethiopia and ORC Macro. Ethiopia Demographic and Health Survey 2016. Addis Ababa: Central Statistical Authority, 2016.

4. Campbell, O.M., et al. Strategies for reducing maternal mortality: getting on with what works. The lancet, 2006, Vol. 368. pp.1284–1299.

5. WHO. Quality, equity, dignity: the network to improve quality of care for maternal, newborn and child health – strategic objectives. Geneva: World Health Organization, 2018.

6. Alkema, L., et al. Global, regional, and national levels and trends in maternal mortality between 1990 and 2015, with scenario-based projections to 2030: a systematic analysis by the UN Maternal Mortality Estimation Inter-Agency Group. The Lancet, 2016, Vol. 387. pp.462–474.

7. Kruk, M.E., et al. High-quality health systems in the Sustainable Development Goals era: time for a revolution. The Lancet Global Health, 2018, Vol. 6. pp.e1196–e1252.

8. Chou, D., et al. Ending preventable maternal and newborn mortality and stillbirths. BMJ, 2015, Vol. 351. p.h4255.

9. Tunçalp, D., et al. Quality of care for pregnant women and newborns—the WHO vision. BJOG: an international journal of obstetrics & gynaecology, 2015, Vol. 122. pp.1045–1049.

10. Sharma, G., et al. Quality care during labour and birth: a multi-country analysis of health system bottlenecks and potential solutions. BMC pregnancy and childbirth, 2015, Vol. 15. p.S2.

11. Lawn, J.E., et al. Stillbirths: Where? When? Why? How to make the data count? The Lancet, 2011, Vol. 377. pp.1448–1463.

12. Sipsma, H., et al. Preferences for home delivery in Ethiopia: provider perspectives. Global public health, 2013, Vol. 8. pp.1014–1026.

13. Nesbitt, R.C., et al. Quality along the continuum: a health facility assessment of intrapartum and postnatal care in Ghana. PloS one, 2013, Vol. 8. p.e81089.

14. Bradley, E.H., et al. Access and quality of rural healthcare: Ethiopian Millennium Rural Initiative. International Journal for Quality in Health Care, 2011, Vol. 23. pp.222–230.

15. Girma, M., et al. Lifesaving emergency obstetric services are inadequate in south-west Ethiopia: a formidable challenge to reducing maternal mortality in Ethiopia. BMC health services research, 2013, Vol. 13. p.459.

16. FMoH of Ethiopia. Ethiopian Hospital Alliance for Quality (EHAQ): Labor & Delivery Change Package for Ethiopian Hospitals. Addis Ababa: FMoH of Ethiopia, 2014.

17. Bayou et al. Structural quality of labor and delivery care in government hospitals of Ethiopia: a descriptive analysis. BMC Pregnancy and Childbirth (2022) 22:523. https://doi.org/10.1186/s12884-022-04850-5.

18. Bradley EH, Curry LA, Ramanadhan S, Rowe L, Nembhard IM, Krumholz HM. Research in action: Using positive deviance to improve quality of health care. Implementation Science 2009; 4:25.

19. Getachew, A., et. al. Quality of care for prevention and management of common maternal and newborn complications: a study of Ethiopia’s hospitals. Baltimore: JHPIEGO, 2011.

20. Yigzaw T., et al. Quality of Midwife-provided Intrapartum Care in Amhara Regional State, Ethiopia. BMC Pregnancy and Childbirth, 2017, Vol. 17. DOI 10.1186/s12884-017-1441-2.

21. Sobel, H.L., et al. Immediate newborn care practices delay thermoregulation and breastfeeding initiation. Acta Paediatrica, 2011, Vol. 100. pp.1127–1133.

22. Miller, S., et al. Quality of care in institutionalized deliveries: the paradox of the Dominican Republic. International Journal of Gynecology & Obstetrics, 2003, Vol. 82. pp.89–103.

23. Smith, H., et al. Implementing the WHO integrated tool to assess quality of care for mothers, newborns and children: results and lessons learnt from five districts in Malawi. BMC pregnancy and childbirth, 2017, Vol. 17. p.271.

24. de Graft-Johnson, J., et al. Cross-sectional observational assessment of quality of newborn care immediately after birth in health facilities across six sub-Saharan African countries. BMJ open, 2017, Vol. 7. p.e014680.

25. Hoque, D.M., et al. An assessment of the quality of care for children in eighteen randomly selected district and sub-district hospitals in Bangladesh. BMC pediatrics, 2012, Vol. 12. p.197.

26. Ogwang, S., et al. Assessment of partogram use during labour in rujumbura health Sub district, Rukungiri district, Uganda. African Health Sciences, 2009, Vol. 9.

27. Kagema, F., et al. Quality of care for prevention and management of common maternal and newborn complications: findings from a National Health Facility Survey in Kenya— are services provided according to international standards. Baltimore: JHPIEGO, 2011.

28. Luwei P, et al. Opportunities for Africa’s Newborns: Practical data, policy and programmatic support for newborn care in Africa. Geneva: World Health Organization, 2014.

29. Worku A.G., et al. Availability and components of maternity services according to providers and users’ perspectives in North Gondar, northwest Ethiopia. Reproductive Health, 2013, Vol. 10.

30. JHPIEGO. Monitoring birth preparedness and complication readiness: tools and indicators for maternal and newborn health. Baltimore : JHPIEGO, 2004.

31. Bowser, D. and Hill, K. Exploring evidence for disrespect and abuse in facility-based childbirth. Boston: USAID-TRAction Project, Harvard School of Public Health, 2010.

32. Leisher, S.H., et al. Quality measurement in family planning: Past present future. Papers from the Bellagio Meeting on Family Planning Quality. Oakland, CA: Metrics for Management, 2016.

33. Mosadeghrad, A.M. Factors influencing healthcare service quality. International journal of health policy and management, 2014, Vol. 3. p.77.

34. Abramson, J. and Abramson, Z.H. Research methods in community medicine: surveys, epidemiological research, programme evaluation, clinical trials. John Wiley & Sons, 2011.

35. The Physician Consortium for Performance Improvement. Measures Development, Methodology, and Oversight Advisory Committee: Recommendations to PCPI Work Groups on Composite Measures. USA: American Medical Association, 2010.

36. Donabedian, A. Evaluating the quality of medical care. The Milbank Quarterly, 2005, Vol. 83. pp.691–729.

37. Bowling, A. Research methods in health: investigating health and health services. UK : McGraw-Hill Education, 2014.

